# The circulating proteome and brain health: Mendelian randomisation and cross-sectional analyses

**DOI:** 10.1101/2023.07.27.23293236

**Authors:** Rosie M. Walker, Michael Chong, Nicolas Perrot, Marie Pigeyre, Danni A. Gadd, Aleks Stolicyn, Liu Shi, MA Archie Campbell, Xueyi Shen, Heather C. Whalley, Alejo Nevado-Holgado, Andrew M. McIntosh, Stefan Heitmeier, Sumathy Rangarajan, Martin O’Donnell, Eric E. Smith, Salim Yusuf, William N. Whiteley, Guillaume Paré

## Abstract

Decline in cognitive function is the most feared aspect of ageing. Poorer midlife cognitive function is associated with increased dementia and stroke risk. The mechanisms underlying variation in cognitive function are uncertain. Here, we assessed associations between 1160 proteins’ plasma levels and two measures of cognitive function, the digit symbol substitution test (DSST) and the Montreal Cognitive Assessment in 1198 PURE-MIND participants. We identified five DSST performance-associated proteins (NCAN, BCAN, CA14, MOG, CDCP1), with NCAN and CDCP1 showing replicated association in an independent cohort, GS (N=1053). MRI-assessed structural brain phenotypes partially mediated (8-19%) associations between NCAN, BCAN, and MOG, and DSST performance. Mendelian randomisation analyses suggested higher CA14 levels might cause larger hippocampal volume and increased stroke risk, whilst higher CDCP1 levels might increase intracranial aneurysm risk. Our findings highlight candidates for further study and the potential for drug repurposing to reduce risk of stroke and cognitive decline.

**Acronyms:** Average causal mediation effect (ACME); brevican (BCAN); carbonic anhydrase 14 (CA14); cluster of differentiation 6 (CD6); CUB-domain containing protein 1 (CDCP1); confidence interval (CI); cerebral microbleed (CMB); cerebrospinal fluid (CSF); digit symbol substitution test (DSST); extracellular matrix (ECM); false discovery rate (FDR); Generation Scotland imaging subsample (GS); Generation Scotland: Scottish Family Health Study (GS:SFHS); instrumental variable (IV); inverse variance weighted (IVW); myelin oligodendrocyte glycoprotein (MOG); Montreal Cognitive Assessment (MoCA); Mendelian randomisation (MR); magnetic resonance imaging (MRI); neurocan (NCAN); perineuronal net (PNN); odds ratio (OR); posterior probability (PP); protein quantitative trait loci (pQTL); pairwise conditional analysis and co-localisation analyses (PWCoCo); Prospective Urban and Rural Epidemiology (PURE); robust adjusted profile score (RAPS); silent brain infarct (SBI); standard deviation (SD); small vessel disease (SVD); white matter hyperintensity (WMH)

## 1. Introduction

Decline in cognitive ability and dementia are the most feared aspects of ageing ^[1]^, providing a strong rationale for investigating the mechanisms underlying cognitive function. Poorer cognitive function is associated with a greater risk of Alzheimer’s dementia and stroke ^[2, 3]^. This may be due to reduced “cognitive reserve”, which postulates that lower premorbid cognitive function leads to worse cognitive impairment for a given degree of neuropathology ^[4]^. Better understanding of these mechanisms could inform strategies for the prevention and treatment of dementia and stroke.

Recent studies have highlighted the potential for investigating cognition and structural brain phenotypes through the study of plasma proteins ^[5–9]^. These studies identified associations between cognitive function and proteins involved in biological functions previously implicated in dementia, including synaptic function, inflammation, immune function, and blood-brain barrier integrity ^[5–7, 9]^. At the time of carrying out this study, previous studies were limited by a focus on a restricted number of proteins and/or purely observational analyses.

Here, we investigated associations between 1160 plasma proteins and cognitive function in the Prospective Urban and Rural Epidemiology (PURE)-MIND cohort ^[10]^, and we sought replication in the independent imaging subsample of the Generation Scotland cohort (henceforth, referred to as “GS”). The proteins assessed represent a wide range of biological processes, permitting a hypothesis-free approach to investigating cognitive function. Subsets of these proteins have been assessed in previous studies, allowing assessment of cross-study replication.

Using a simple measure of processing speed, the digit symbol substitution task (DSST), and a cognitive screening tool, the Montreal Cognitive Assessment (MoCA), we carried out a screen for cognition-associated proteins, and then employed mediation analyses to assess the proportion of the protein expression-cognition relationship that could be explained by structural brain phenotypes, including measures of brain volume and white matter hyperintensity (WMH) volume. WMH is an MRI marker of white matter damage, and is one of the manifestations of age-related cerebral small vessel disease. Two sample Mendelian randomisation (MR) analyses were performed to assess potentially causal effects of genetically predicted protein levels on genetically predicted cognitive function, brain structure, stroke subtypes, and Alzheimer’s disease (see **Figure 1** for an overview of the study design).

**Figure 1.**
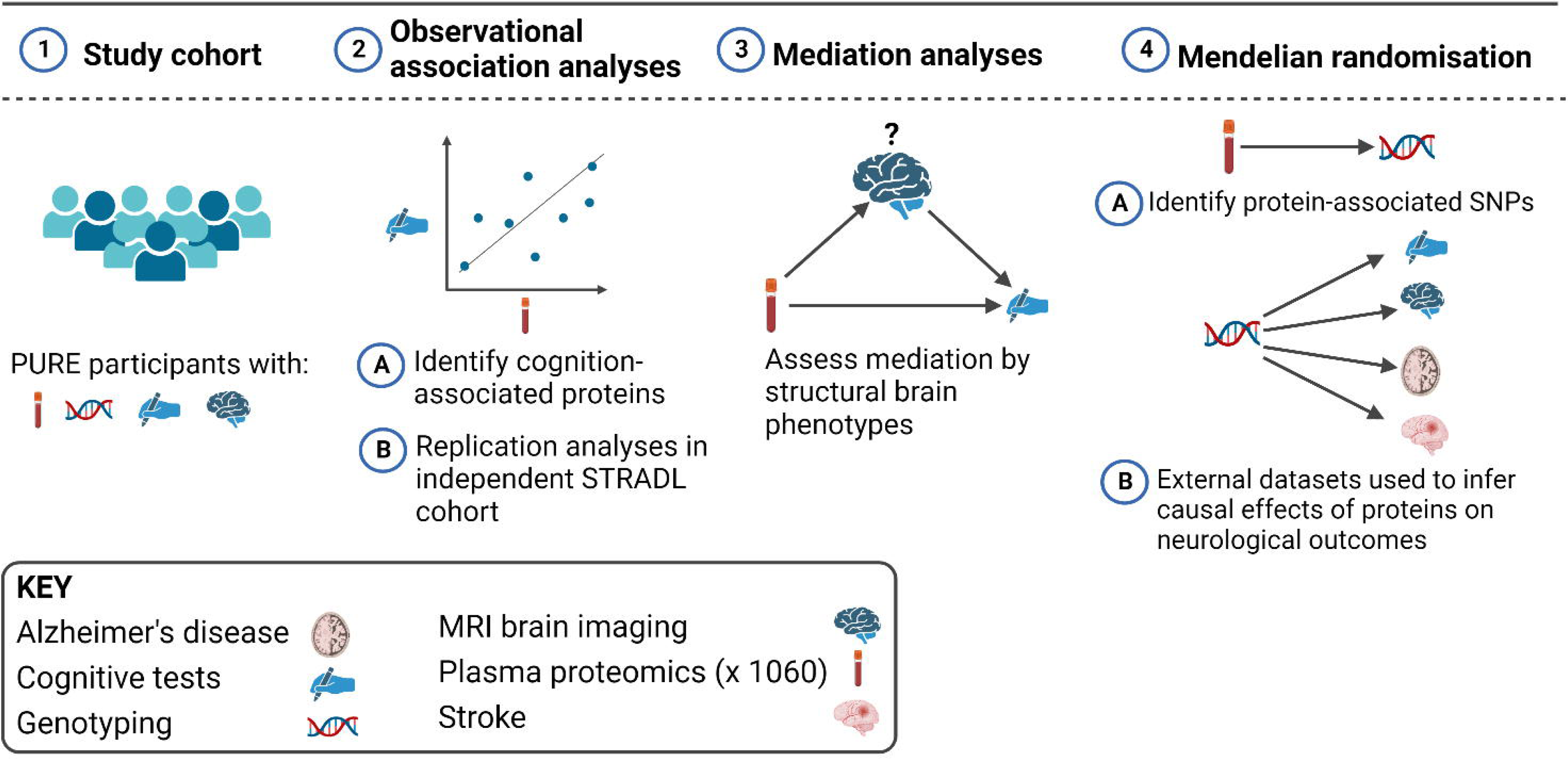
Overview of the study design. This study involved European (N = 3514), Latin (N = 4309), and Persian (N = 1332) PURE participants for whom genetic and plasma proteomic data were available. Observational analyses to detect plasma biomarkers of cognitive function were performed in the subset of these participants who were enrolled in the PURE-MIND sub-study (N = 1198), for whom plasma protein (N = 1060 proteins) and MRI measurements were available. Mediation analyses were performed to assess whether any observed associations between protein levels and cognitive function were mediated by structural brain phenotypes ascertained by MRI. Finally, two-sample Mendelian randomisation analyses were performed to assess potentially causal effects of genetically-predicted cognition-associated protein levels on genetically-predicted neurological outcomes. For these analyses, genetic instrumental variables for protein levels were identified in the European, Latin, and Persian PURE participants, and associations with neurological outcomes were assessed using external (non-PURE) datasets.

## 2. Results

### Cohort characteristics

Key demographic, cognitive, brain MRI, and health variables for the participants in PURE-MIND (N=1198) and GS (N=1053) are summarised in **Table 1**. Participants in PURE-MIND were significantly younger than participants in GS (PURE-MIND: mean = 54.5 years (SD = 8.05 years); GS: mean = 59.9 years (SD = 9.59 years); *p* < 2.2 x 10^−16^), and a small, but significant, between-cohort difference in DSST score was observed (PURE-MIND: mean = 69.5 (SD = 15.3); GS: mean = 68.1 (SD = 15.2); *p* = 0.0298). Differences in the levels/types of education received by the two cohorts were observed (*p* = 5 x 10^−4^). The two cohorts also differed significantly on several brain volume measurements: PURE-MIND participants have a smaller total brain volume (PURE-MIND: mean = 1058 cm^3^ (SD = 110 cm^3^); GS: mean = 1069 cm^3^ (SD = 109 cm^3^); *p* = 0.0212), cerebral white matter volume (PURE-MIND: mean = 447 cm^3^ (SD = 60.5 cm^3^); GS: mean = 455 cm^3^ (SD = 56.9 cm^3^); *p* = 1.87 x 10^−3^), and hippocampal volume (PURE-MIND: mean = 3.92 cm^3^ (SD = 0.433 cm^3^); GS: mean = 4.18 cm^3^ (SD = 0.437 cm^3^); *p* = < 2.2 x 10-^16^) than GS participants, whilst GS participants have a smaller intracranial volume (ICV) than PURE-MIND participants (PURE-MIND: mean = 1491 cm^3^ (SD = 159 cm^3^); GS: mean = 1400 cm^3^ (SD = 225 cm^3^); *p* = < 2.2 x 10-^16^). The two cohorts also differed significantly on other health-related variables that were not directly assessed in this study (**Table 1**).

**Table 1.**
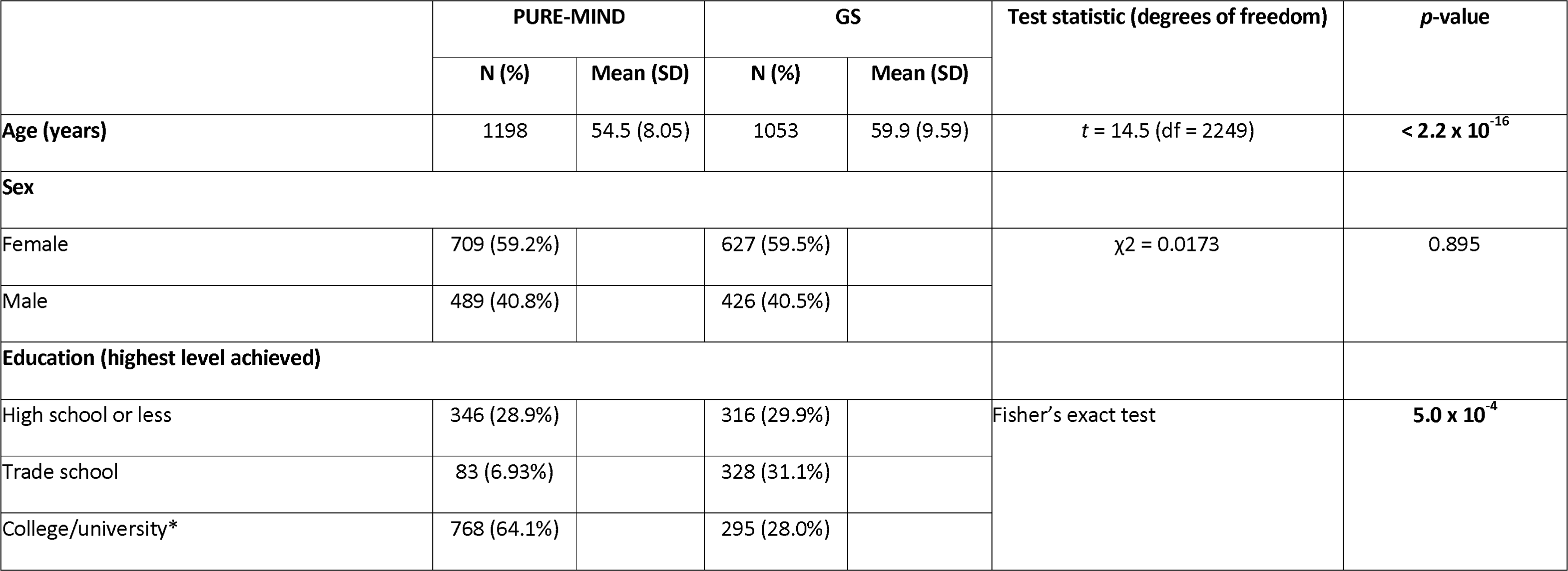
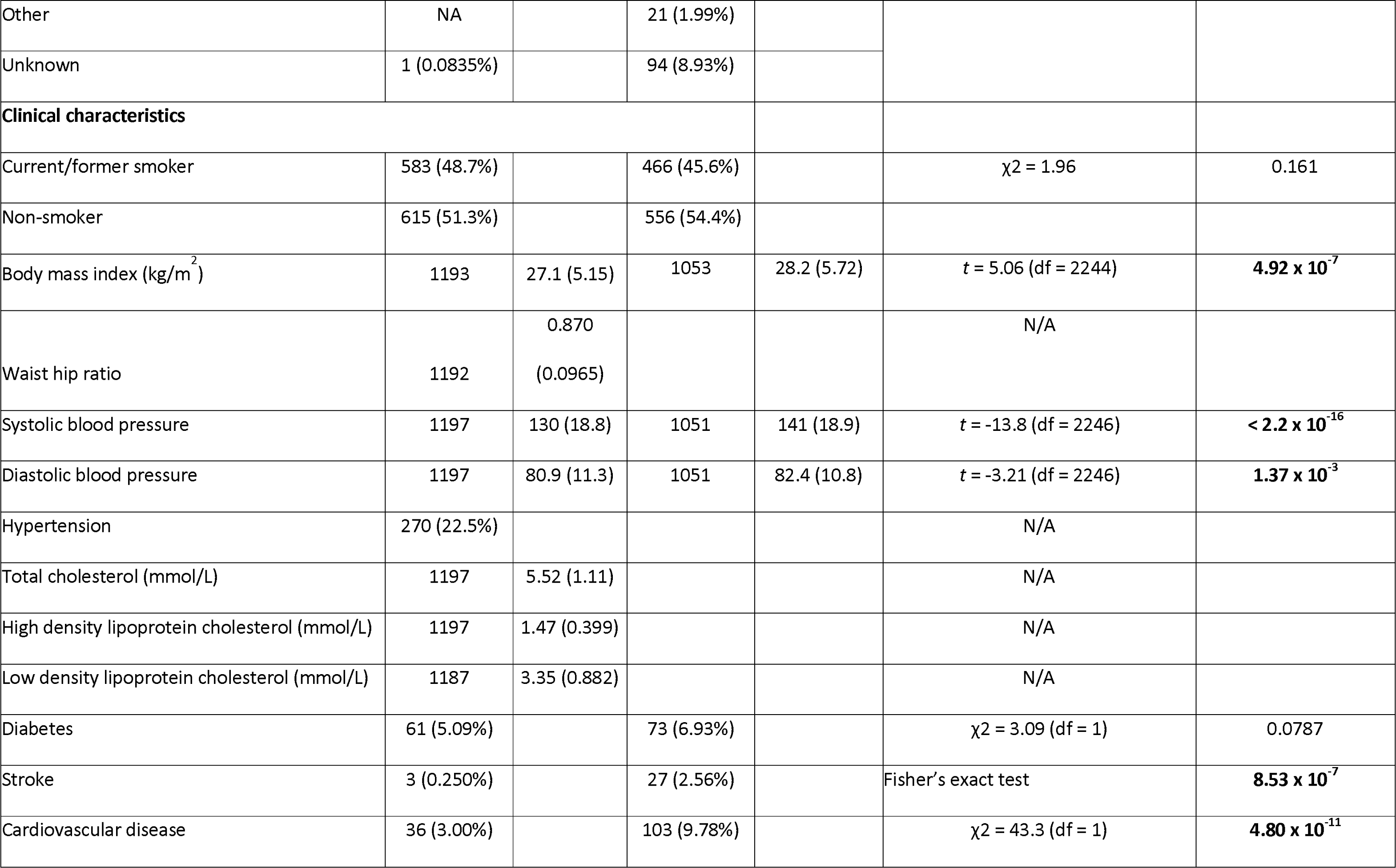
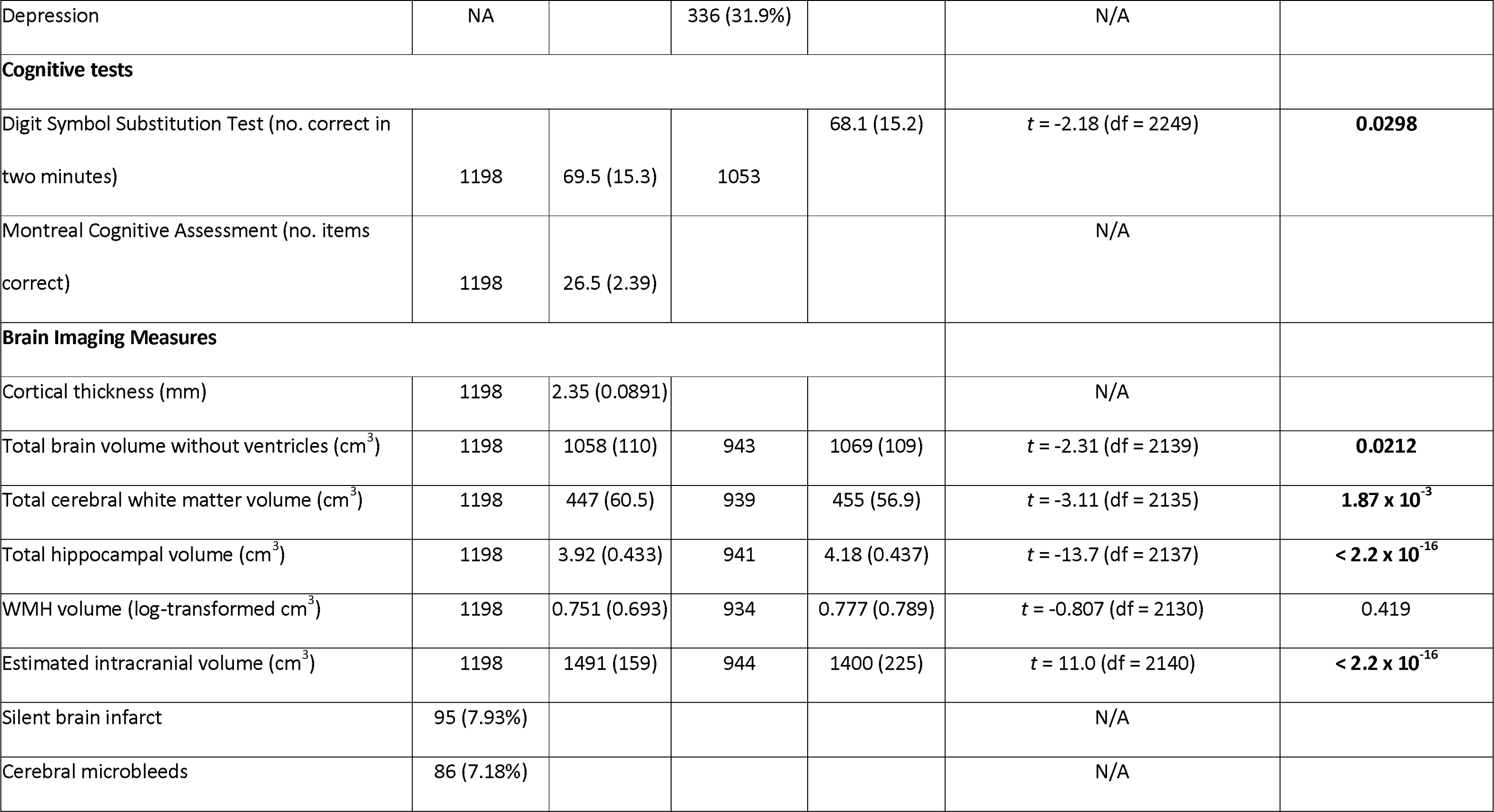
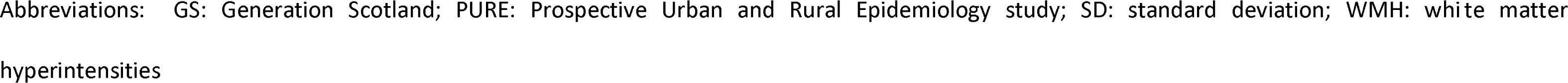
Demographic information for the discovery sample (PURE-MIND) and the replication sample (GS). Basic sample demographic information is presented together with information on relevant clinical characteristics, cognitive performance, and structural brain imaging measures. The number of participants for whom information was available for each variable are indicated in the “N” columns. In GS, there were participants with missing information for smoking (N = 31), hypertension (N = 2), diabetes (N = 1), stroke (N = 1), and depression (N = 2). *PURE-MIND: college or university; GS: university. For variables that were measured in both cohorts, a statistical comparison was performed and significant _p_-values (_p_ < 0.05) are indicated in bold.

|  | PURE-MIND |  | GS |  | Test statistic (degrees of freedom) | *p*-value |
| --- | --- | --- | --- | --- | --- | --- |
|  | N (%) | Mean (SD) | N (%) | Mean (SD) |  |  |
| **Age (years)** | 1198 | 54.5 (8.05) | 1053 | 59.9 (9.59) | *t* = 14.5 (df = 2249) | **< 2.2 x 10<sup>-16</sup>** |
| **Sex** |  |  |  |  |  |  |
| Female | 709 (59.2%) | | 627 (59.5%) | | $\chi^2 = 0.0173$ | 0.895 |
| Male | 489 (40.8%) |  | 426 (40.5%) |  |  |  |
| **Education (highest level achieved)** |  |  |  |  |  |  |
| High school or less | 346 (28.9%) |  | 316 (29.9%) |  | Fisher's exact test | **5.0 x 10<sup>-4</sup>** |
| Trade school | 83 (6.93%) |  | 328 (31.1%) |  |  |  |
| College/university* | 768 (64.1%) |  | 295 (28.0%) |  |  |  |
| Other | NA |  | 21 (1.99%) |  |  |  |
| Unknown | 1 (0.0835%) |  | 94 (8.93%) |  |  |  |
| **Clinical characteristics** |  |  |  |  |  |  |
| Current/former smoker | 583 (48.7%) | | 466 (45.6%) | | $\chi^2 = 1.96$ | 0.161 |
| Non-smoker | 615 (51.3%) |  | 556 (54.4%) |  |  |  |
| Body mass index (kg/m<sup>2</sup>) | 1193 | 27.1 (5.15) | 1053 | 28.2 (5.72) | $t = 5.06$ (df = 2244) | **$4.92 \times 10^{-7}$** |
| Waist hip ratio | 1192 | 0.870<br>(0.0965) |  |  | N/A |  |
| Systolic blood pressure | 1197 | 130 (18.8) | 1051 | 141 (18.9) | $t = -13.8$ (df = 2246) | **$< 2.2 \times 10^{-16}$** |
| Diastolic blood pressure | 1197 | 80.9 (11.3) | 1051 | 82.4 (10.8) | $t = -3.21$ (df = 2246) | **$1.37 \times 10^{-3}$** |
| Hypertension | 270 (22.5%) |  |  |  | N/A |  |
| Total cholesterol (mmol/L) | 1197 | 5.52 (1.11) |  |  | N/A |  |
| High density lipoprotein cholesterol (mmol/L) | 1197 | 1.47 (0.399) |  |  | N/A |  |
| Low density lipoprotein cholesterol (mmol/L) | 1187 | 3.35 (0.882) |  |  | N/A |  |
| Diabetes | 61 (5.09%) | | 73 (6.93%) | | $\chi^2 = 3.09$ (df = 1) | 0.0787 |
| Stroke | 3 (0.250%) |  | 27 (2.56%) |  | Fisher's exact test | **$8.53 \times 10^{-7}$** |
| Cardiovascular disease | 36 (3.00%) | | 103 (9.78%) | | $\chi^2 = 43.3$ (df = 1) | **$4.80 \times 10^{-11}$** |
| Depression | NA |  | 336 (31.9%) |  | N/A |  |
| **Cognitive tests** |  |  |  |  |  |  |
| Digit Symbol Substitution Test (no. correct in two minutes) | 1198 | 69.5 (15.3) | 1053 | 68.1 (15.2) | $t = -2.18$ (df = 2249) | **0.0298** |
| Montreal Cognitive Assessment (no. items correct) | 1198 | 26.5 (2.39) |  |  | N/A |  |
| **Brain Imaging Measures** |  |  |  |  |  |  |
| Cortical thickness (mm) | 1198 | 2.35 (0.0891) |  |  | N/A |  |
| Total brain volume without ventricles (cm<sup>3</sup>) | 1198 | 1058 (110) | 943 | 1069 (109) | $t = -2.31$ (df = 2139) | **0.0212** |
| Total cerebral white matter volume (cm<sup>3</sup>) | 1198 | 447 (60.5) | 939 | 455 (56.9) | $t = -3.11$ (df = 2135) | **$1.87 \times 10^{-3}$** |
| Total hippocampal volume (cm<sup>3</sup>) | 1198 | 3.92 (0.433) | 941 | 4.18 (0.437) | $t = -13.7$ (df = 2137) | **$< 2.2 \times 10^{-16}$** |
| WMH volume (log-transformed cm<sup>3</sup>) | 1198 | 0.751 (0.693) | 934 | 0.777 (0.789) | $t = -0.807$ (df = 2130) | 0.419 |
| Estimated intracranial volume (cm<sup>3</sup>) | 1198 | 1491 (159) | 944 | 1400 (225) | $t = 11.0$ (df = 2140) | **$< 2.2 \times 10^{-16}$** |
| Silent brain infarct | 95 (7.93%) |  |  |  | N/A |  |
| Cerebral microbleeds | 86 (7.18%) |  |  |  | N/A |  |

Abbreviations: GS: Generation Scotland; PURE: Prospective Urban and Rural Epidemiology study; SD: standard deviation; WMH: white matter hyperintensities

Scores on the DSST were normally distributed in both PURE-MIND and GS (**Supplementary Figure 1**), but scores on the MoCA showed a leftward skew in PURE-MIND (**Supplementary Figure 1**).

### Identification of protein biomarkers of cognitive function and enrichment analyses

Five proteins were associated with DSST performance in PURE-MIND (**Figure 2; Figure 3; Supplementary Table 1; Supplementary Figure 2**). Higher plasma levels of neurocan (NCAN; β = 2.03 (indicating a 2.03 higher DSST score per a standard deviation higher NCAN level), *p* = 9.11 x 10^−8^), brevican (BCAN; β = 1.91, *p* = 5.56 x 10^−7^), carbonic anhydrase 14 (CA14; β = 1.90, *p* = 5.90 x 10^−7^), and myelin-oligodendrocyte glycoprotein (MOG; β = 1.82, *p* = 2.29 x 10^−6^), and lower levels of CUB domain-containing protein 1 (CDCP1; β = −1.57, *p* = 3.97 x 10^−5^) were associated with significantly better DSST performance. Adjustment for educational attainment modestly attenuated the effect estimate for all five proteins (**Supplementary Table 1**). Levels of NCAN, BCAN and MOG were positively correlated (0.251 ≤ *r* ≥ 0.615; all *p* < 2.20 x 10^−16^), whilst CDCP1 and CA14 expression levels were negatively correlated (*r* = −1.01, *p* = 4.82 x 10^−4^; **Supplementary Table 2**). Three proteins (NCAN, BCAN and CDCP1) proteins were also measured in GS, of which two replicated their association with DSST performance: NCAN (β = 1.40, *p* = 1.07 x 10^−3^) and CDCP1 (β = −1.99, *p* = 9.21 x 10^−6^; **Figure 3**). MoCA performance was not associated with the level of any protein (all *p* ≥ 2.34 x 10^−4^; **Supplementary Table 3; Supplementary Figure 3**). When considering the 129 proteins that were nominally significantly associated (*p* < 0.05) with DSST score in PURE-MIND and measured in GS (**Supplementary Table 4**), their effect estimates showed a strong, statistically significant between-cohort correlation (*r* = 0.625, *p* = 2.34 x 10^−15^).

**Figure 2.**
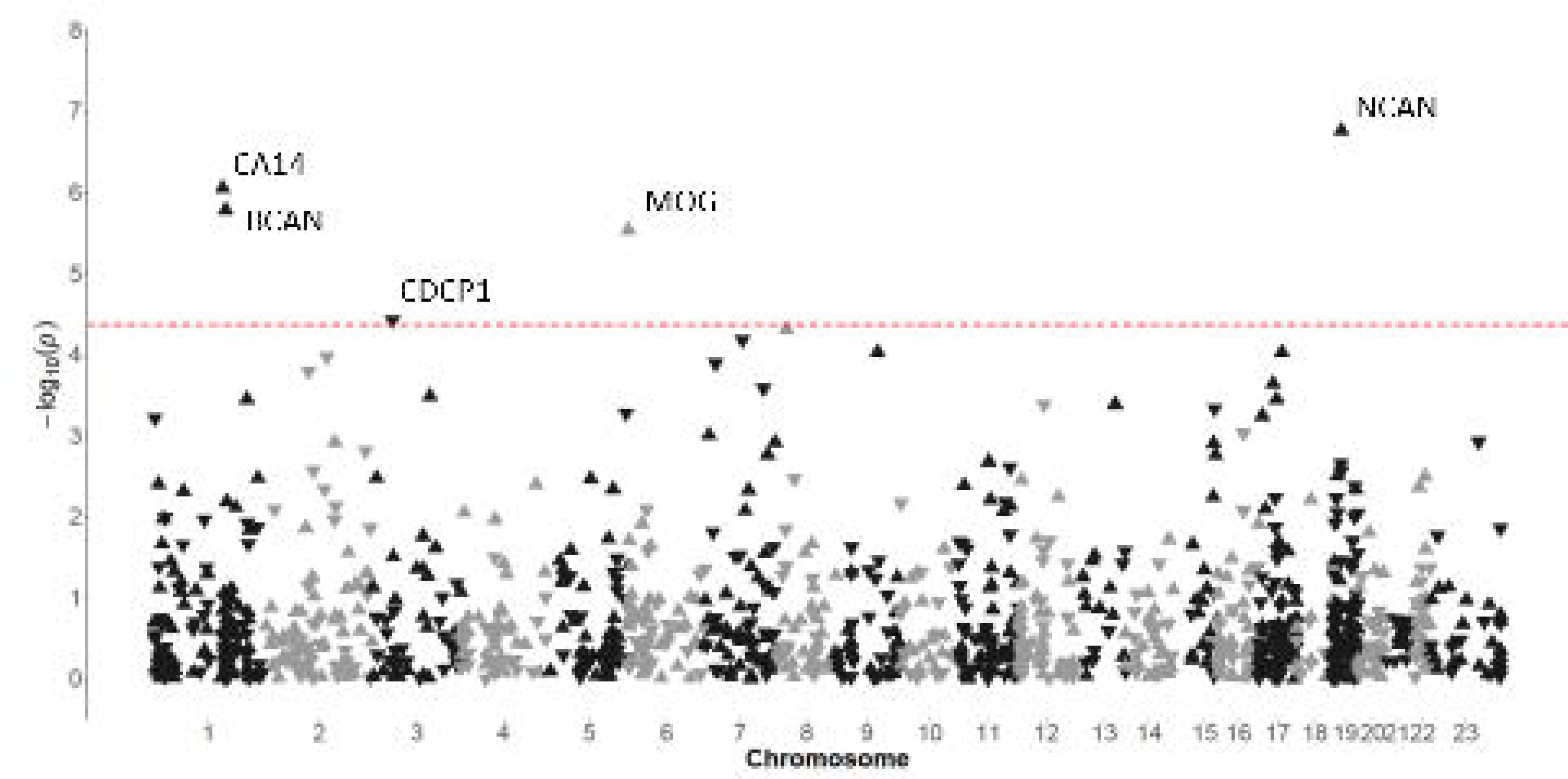
Manhattan plot indicating associations between the levels of plasma proteins and performance on the DSST in participants from the PURE-MIND cohort (N = 1198). Each protein is represented by a triangle with upwards-facing triangles indicating a positive association with DSST performance and downwards-facing triangles indicating a negative association with DSST performance. The position of each protein on the x-axis is determined by the genomic location of its corresponding gene and the position on the y-axis is determined by the –log_10_ *p*-value. The dashed horizontal line indicates the Bonferroni-corrected significance threshold (*p* = 4.31 x 10^−5^) required to maintain a 5% type I error rate.

**Figure 3.**
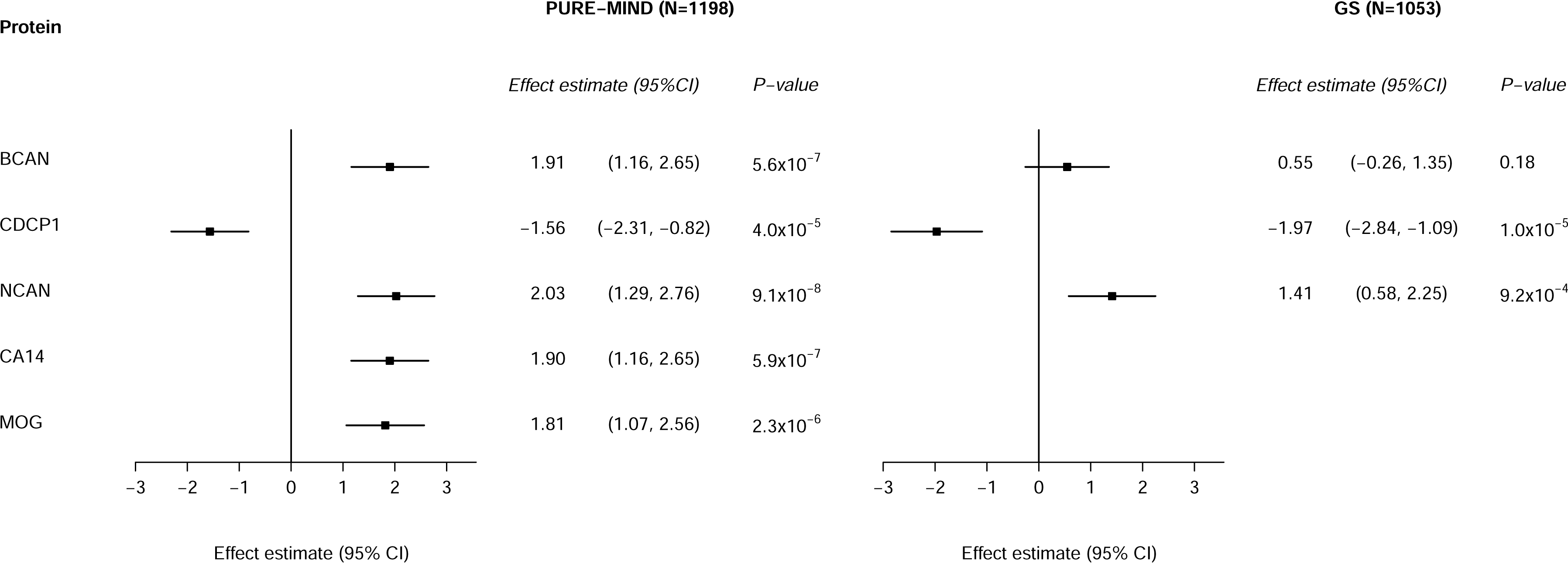
Forest plot indicating the association between protein levels and DSST performance for significantly associated proteins. For each protein, the difference in DSST score associated with a standard deviation higher level of protein is shown, together with the 95% confidence interval. Abbreviations: BCAN: brevican; CA14: carbonic anhydrase 14; CDCP1: CUB-domain containing protein 1; CI: confidence interval; GS: Generation Scotland imaging subsample; MOG: myelin oligodendrocyte glycoprotein; NCAN: neurocan; PURE: Prospective Urban and Rural Epidemiology study.

Proteins nominally associated (*p* < 0.05) with DSST performance (N = 184) were enriched for brain-expressed proteins, most significantly for proteins with hippocampal expression (FDR-corrected *p* = 0.0154; **Supplementary Table 5**). Better DSST performance was nominally associated with lower levels of 90 proteins. These proteins mapped to the following immune pathways “interleukin-10 signalling”, “glomerulonephritis”, “regulation of granulocyte chemotaxis”, “positive regulation of leukocyte chemotaxis”, “positive regulation of leukocyte migration”, and “inflammation” (FDR-corrected *p* ≤ 0.0337; **Supplementary Table 6**).

### Structural brain phenotypes as mediators of protein biomarker-DSST performance associations

In PURE-MIND, better DSST performance was associated with greater cerebral white matter volume (β = 0.0615, *p* = 4.34 x 10^−7^), greater total brain volume (β = 0.0349, *p* = 9.64 x 10^−6^), greater hippocampal volume (β = 2.97, *p* = 4.79 x 10^−3^), and lower log-transformed WMH volume (β = −3.20, *p* = 1.18 x 10^−6^). These associations replicated in GS (**Supplementary Table 7**).

Assessment of the relationships between protein levels and DSST-associated structural brain phenotypes in PURE-MIND revealed systematic differences between those proteins for which higher levels were associated with better DSST performance (NCAN, BCAN, CA14, and MOG), and CDCP1, which was negatively associated with DSST performance (**Figure 4**). Whilst NCAN, BCAN, CA14, and MOG showed a positive direction of association with total brain, cerebral white matter, and hippocampal volume measurements and a negative association with WMH volume, the converse was true for CDCP1. The associations between NCAN levels and total brain, cerebral white matter, and hippocampal volumes reached statistical significance (*p* ≤ 2.56 x 10^−5^) and were replicated in GS (*p* ≤ 6.70 x 10^−3^). BCAN levels were significantly associated with all four brain volumes (*p* ≤ 4.36 x 10^−4^), with the associations with total brain and cerebral white matter volumes replicating in GS (*p* ≤ 44 x 10^−3^). The associations between MOG levels and total brain and cerebral white matter volumes attained statistical significance (*p* ≤ 2.63 x 10^−9^), but could not be assessed in GS. We did not identify any significant associations with CA14 or CDCP1 levels after multiple testing correction.

**Figure 4.**
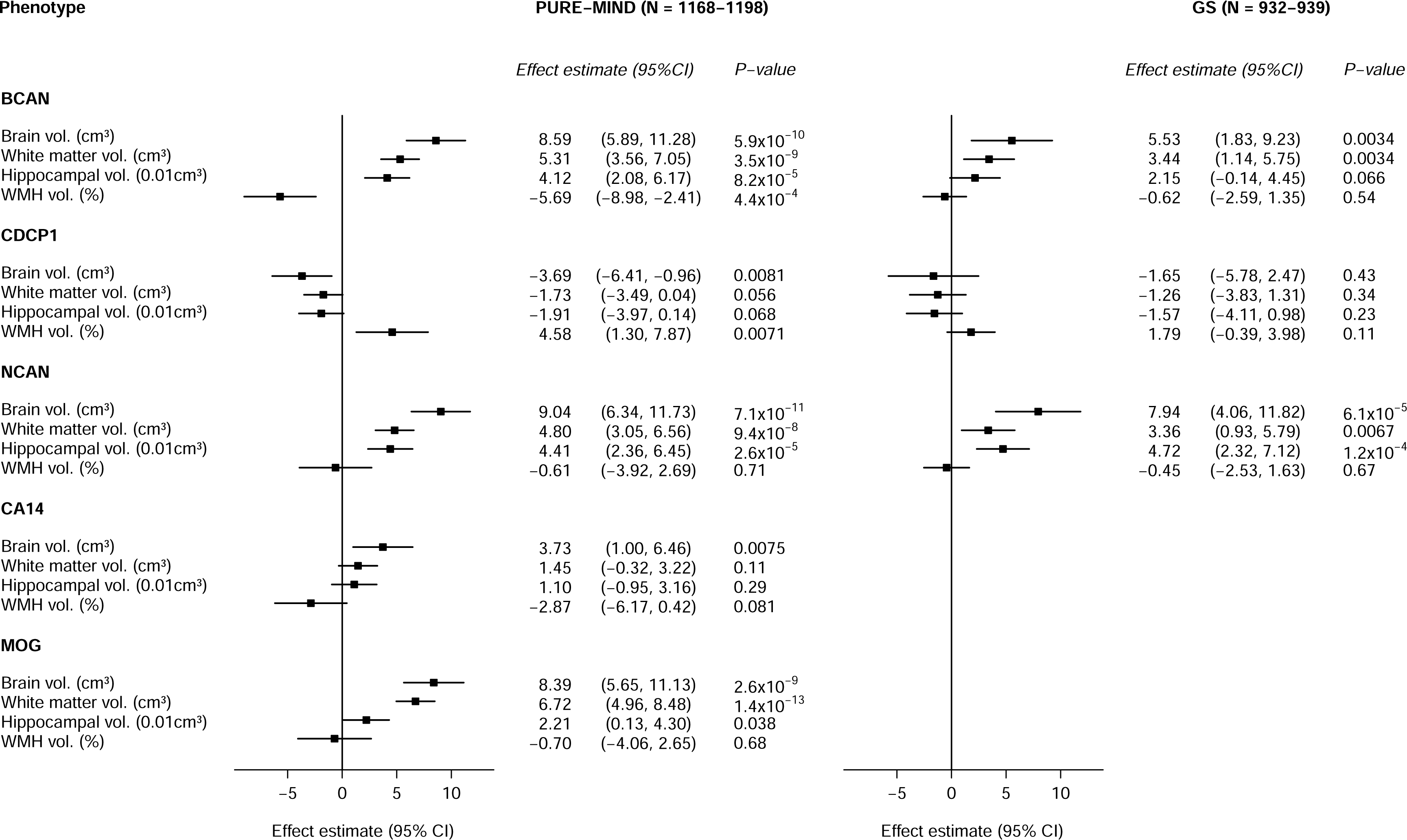
Forest plots indicating the association between the levels of DSST-associated proteins and DSST-associated structural brain phenotype. For each protein, the effect estimate (change in brain volume (cm^3^ or %) per standard deviation increase in protein expression) is shown, together with the 95% confidence interval. Abbreviations: BCAN: brevican; CA14: carbonic anhydrase 14; CDCP1: CUB-domain containing protein 1; CI: confidence interval; GS: Generation Scotland imaging subsample; MOG: myelin oligodendrocyte glycoprotein; NCAN: neurocan; PURE: Prospective Urban and Rural Epidemiology study; WMH: white matter hyperintensity.

In PURE-MIND, cerebral white matter volume explained a significant proportion of variance in the relationship between MOG (19.2%), BCAN (14.9%), and NCAN (12.7%) levels and DSST performance (all *p* < 2 x 10^−16^) (**Supplementary Table 8**). After controlling for cerebral white matter volume, the average effect estimates (based on 1000 bootstrap resamples) for these proteins were reduced from 1.81 to 1.47 (MOG), 1.91 to 1.62 (BCAN), and 2.03 to 1.77 (NCAN). Log-transformed WMH volume was a significant partial mediator of the association between BCAN levels and DSST performance (*p* = 0.002). Controlling for log-transformed WMH resulted in a reduction in the effect estimate from 1.91 to 1.75 (8% mediation).

### Identification of potentially causal relationships between protein levels and cognitive function, structural brain phenotypes, and disease outcomes

Inverse variance weighted (IVW) MR analyses were performed to assess the effects of genetically predicted CA14, CDCP1, and MOG levels on cognitive function, structural brain phenotypes, and Alzheimer’s disease, and stroke. Protein quantitative trait loci (pQTLs) located in *cis* to the genes encoding the proteins-of-interest acted as instrumental variables (IVs) for plasma protein levels (**Supplementary Table 9**). An insufficient number of pQTLs precluded the assessment of BCAN andNCAN with any of the outcomes-of-interest. For MOG, limited overlap between the pQTLs and SNPs included in the outcome GWASs meant only a subset of the outcomes-of-interest could be assessed.

A one standard deviation higher level of genetically predicted plasma CA14 was associated with a larger hippocampal volume (β = 0.0971 [95% CI: 0.0300 to 0.164], *p* = 4.58 x 10^−3^), and a greater risk of all stroke (odds ratio (OR) = 1.08 [95% CI: 1.02 to 1.14], *p* = 6.97 x 10^−3^; **Supplementary Table 10**). A one standard deviation higher level of genetically predicted plasma CDCP1 was associated with an increased risk of intracranial aneurysm (OR = 1.22 [95% CI: 1.02 to 1.47], *p* = 0.0280). These associations were corroborated by similar effect estimates from weighted median and MR-RAPS analyses. No evidence of directional or horizontal pleiotropy were observed, and the correct causal direction was assessed. No significant associations were observed between the genetically predicted levels of CA14 or CDCP1 and risk of Alzheimer’s disease (*p* ≥ 0.125) (NB. A lack of significant pQTLs precluded the assessment of BCAN, MOG, or NCAN).

Sensitivity analyses were performed in which instrumental variables (IVs) were selected using a stricter threshold for independence. For genetically predicted CA14, these analyses supported the association with hippocampal volume (β = 0.144 [95% CI: 0.0435 to 0.244], *p* = 4.97 x 10^−3^), and produced a consistent, although non-significant, effect estimate for the association with risk for all stroke (**Supplementary Table 10**). For CDCP1, the sensitivity analyses identified a consistent, although non-significant, effect estimate for the association with risk for intracranial aneurysm.

To assess whether between-population heterogeneity in pQTL effects could have affected our findings, we sought to replication using random effects meta-analysis (**Supplementary Table 10**). For genetically predicted CA14, this approach supported the significant associations with hippocampal volume (β = 0.0953 [95% CI: 0.0586 to 0.132], *p* = 3.63 x 10^−7^), and risk for all stroke (OR = 1.06 [95% CI: 1.03 to 1.09], *p* = 2.56 x 10^−4^). For genetically predicted CDCP1, it was only possible to assess the association with risk for intracranial aneurysm using a single IV; this identified a significant association (OR = 1.32 [95% CI: 1.08 to 1.61], *p* = 6.93 x 10^−3^).

Pairwise Conditional Analysis and Co-localisation Analyses (PWCoCo) were performed to assess the presence of a shared variant for each of the five proteins-of-interest and the same outcomes as assessed by two-sample MR analyses. We were only adequately powered to assess co-localisation between SNPs associated with one pair of traits: MOG plasma level and cognitive function. We did not observe any evidence in support of co-localisation or conditional co-localisation (posterior probability (PP)4/PP3 ≤ 4.81 x 10^−4^).

## 3. Discussion

In this large-scale analysis of the associations between the plasma levels of 1160 proteins and cognitive function, we identify CA14 and CDCP1 as being associated with processing speed, as measured by the DSST, and having potentially causal effects on hippocampal volume and stroke (CA14)and intracranial aneurysm (CDCP1).

Other proteins (BCAN, NCAN, and MOG) were associated with DSST performance and important structural brain phenotypes, with cerebral white matter volume mediating a significant proportion (13-19%) of the relationship between the levels of all three proteins and DSST performance, and WMH volume mediating 8% of the relationship between BCAN levels and DSST performance. A lack of genetic instruments precluded the assessment of potentially causal effects of BCAN and NCAN with any outcome-of-interest, and MOG with several outcomes-of-interest.. Enrichment analyses of proteins that were nominally significantly associated with DSST performance revealed a significant enrichment for brain-expressed proteins.

There were no significant associations between plasma protein levels and performance on the MoCA. This might reflect the fact that the MoCA is a screening tool for mild cognitive impairment ^[11]^, meaning its sensitivity to detect variation in cognitive function in non-clinical groups is likely to be limited. The maximum MoCA score is 30 and scores higher than 26 indicate normal function. A high mean score (26.5) and a left-skewed distribution indicate a ceiling effect, which likely limited power to detect associations between protein levels and MoCA score in PURE-MIND.

CA14 is one of fifteen isoforms of the carbonic anhydrase family of zinc metalloprotease enzymes, which catalyse the reversible hydration of carbon dioxide ^[12]^. CA14 is expressed by neurons ^[13]^ and involved in regulating extracellular pH following synaptic transmission ^[14, 15]^. Consistent with our findings, acute inhibition of CA14 leads to impaired performance on cognitive tasks in mice ^[16]^. Carbonic anhydrase activation may lead to beneficial cognitive effects in rodents ^[17]^. In keeping with our MR results, there are neuroprotective effects of carbonic anhydrase inhibition in models of amyloidosis, Huntington’s disease, and ischaemic and haemorrhagic stroke ^[17]^. The mechanisms by which carbonic anhydrase inhibition and activation exert their effects are uncertain ^[16, 17]^. FDA-approved carbonic anhydrase inhibitors, and thus the majority of carbonic anhydrase inhibitors investigated to date, are pan-carbonic anhydrase inhibitors. Of the carbonic anhydrase family members measured in our study (CA1, 2, 3, 4, 5A, 6, 9, 12, 13, and 14), only CA14 levels were significantly associated with DSST performance. Further studies are required to determine the therapeutic potential for carbonic anhydrase modulation in the context of cognitive impairment, Alzheimer’s disease, and stroke.

The extracellular matrix (ECM) proteins NCAN and BCAN are brain-specific chondroitin sulfate proteoglycans, which are expressed by neurons and astrocytes (NCAN and BCAN), and oligodendrocytes (BCAN). They contribute to the formation of a specialised structure, the perineuronal net (PNN), which plays a key role in memory and neuronal plasticity, and which is disrupted in Alzheimer’s disease ^[18]^. Our findings are consistent with those of Harris et al. (2020) ^[5]^, who found plasma levels of NCAN and BCAN were positively associated with brain volume. Plasma levels of both NCAN and BCAN have previously been shown to be positively associated with general cognitive function and DSST performance ^[9]^, whilst BCAN levels have been found to be positively associated with Mini Mental State Examination performance and reduced in patients with Alzheimer’s disease or mild cognitive impairment ^[7]^. Mice that are lacking either NCAN or BCAN expression show normal development and memory function but reduced hippocampal long term potentiation ^[19, 20]^, whilst quadruple knock-outs, which lack NCAN, BCAN, and two additional ECM proteins (tenascin-C and tenascin-R) show an altered ratio of excitatory to inhibitory synapses and a reduction in the number and complexity of hippocampal PNNs ^[21]^. Genetic variation in the gene encoding A Disintegrin and Metalloproteinase with Thrombospondin Motifs 4 (ADAMTS4), which degrades the four members of the lectican family (including NCAN and BCAN), has been implicated in Alzheimer’s disease ^[22]^. Taken together, the evidence suggests NCAN, BCAN and their regulators as molecules-of-interest in Alzheimer’s disease.

MOG is an oligodendrocyte-expressed membrane glycoprotein, the exact function of which is unknown ^[23]^. CDCP1 is a widely expressed transmembrane glycoprotein that acts as a ligand for T cell-expressed Cluster of Differentiation 6 (CD6), and is implicated in autoimmune conditions ^[24]^. CDCP1 is amenable to modulation by approved drug treatments: Itolizumab, which is used to treat psoriasis, disrupts CDCP1-CD6 binding and downregulates T-cell-mediated inflammation ^[25]^, whilst atomoxetine, a treatment for attention deficit hyperactivity disorder, which is being considered for the treatment of mild cognitive impairment, reduced cerebrospinal fluid (CSF) CDCP1 levels ^[26]^. Intriguingly, findings in mice suggest a functional link between CDCP1 and MOG ^[6]^.

Our study has several strengths. We measured 1160 proteins, associated with a wide range of physiological processes, in a large, well-characterised cohort. Replication analyses, where possible, were performed in an independent cohort in which proteins were measured using an independent methodology. The availability of genetic and brain MRI data permitted an exploration of causality and putative causal pathways. The use of MR to identify potentially causal associations will have offered protection against some of the common confounders of observational analyses ^[27]^, with the use of multiple MR methods, which generally gave concordant estimates of effect, mitigating against the individual biases of different MR methodologies ^[28]^. Moreover, by requiring instrumental variables to be located in *cis* to their target protein, we limited the chance of pleiotropic effects ^[29]^.

There are also several limitations to consider.

First, the 1160 proteins measured represent a small subset of the circulating proteome ^[30]^. Although these proteins are involved in a wide range of biological functions represented by all 13 Olink Target 96 panels, limitations to our understanding of the proteome mean that is not possible to assess the extent to which these proteins are representative of the rest of the proteome. Replication analyses were only performed for those proteins for which data were available in the GS cohort, meaning that we did not assess replication of CA14 or MOG.

Second, the availability of suitable IVs mean that our primary MR analyses were only performed for CA14, CDCP1, and MOG. Whilst we required a minimum of three IVs for the primary MR analyses, our sensitivity analyses, in which a stricter threshold for independence was applied to the IVs necessitated the use of fewer than three IVs in each analysis. As such, the results of the sensitivity analyses should be interpreted with this caveat in mind.

Third, for all but one pair of traits, we were insufficiently powered to assess co-localisation between genetic variants associated with protein level and cognition, structural brain phenotypes, and disease outcomes. This means that it is possible that significant MR findings might reflect the presence of separate causal variants in linkage disequilibrium (LD) with one another ^[31]^

Fourth, we measured protein levels in the plasma, rather than in the brain or CSF. It is, however, important to note the striking enrichment for brain-expressed proteins amongst the DSST-associated proteins. Previous analyses of the GS cohort, in which replication was sought in the present study, have identified the levels of several plasma proteins as being associated with multiple markers of brain health ^[8]^. These findings support the use of the plasma to assess brain-related phenotypes and emphasise the need for additional research to explain the mechanisms controlling the efflux of brain-expressed proteins into the bloodstream in non-clinical populations. Moreover, the use of *cis* pQTLs, which are likely to be shared across tissues ^[32]^, as IVs in our MR analyses, supports the possibility that the MR-identified associations reflect the actions of the proteins-of-interest in the brain.

In summary, we identified protein biomarkers of cognitive function that may causally affect brain structure and risk for stroke and intracranial aneurysm. Notwithstanding the need for replication, our findings prompt several hypotheses that should be assessed by future studies. Our apparently paradoxical findings of higher CA14 levels being associated with both better cognitive function and increased stroke risk suggest that molecular findings can inform a more nuanced understanding of the relationship between premorbid cognitive function and neurological disease risk. It is possible that improved risk stratification may be achieved through the combination of cognitive assessment and biomarker measurement. The availability of approved drugs targeting our identified proteins raises the possibility of drug repurposing for novel therapeutic interventions to prevent cognitive decline, stroke, and intracranial aneurysm.

## 4. Methods

### Sample information

This study used data from participants of self-reported European (N = 3514), Latin (N = 4309), or Persian (N = 1332) ancestry from the Population Urban Rural Epidemiology (PURE) biomarker sub-study ^[33]^ (**Supplementary Information**). African (n=659), South Asian (n=604), East Asian (n=314), and Arab (n=204) participants were excluded to align PURE genetic data with external genetic datasets, which are predominantly European. Participants of Latin and Persian ancestry were included due to their genetic overlap with European participants ^[33]^.

The PURE biomarker study also included European participants enrolled in PURE-MIND (N = 1198) ^[10]^ (**Supplementary Information**).

The European, Latin, and Persian PURE biomarker cohort participants were used to identify protein biomarker pQTLs ^[33]^, for use in MR analyses, whilst data from the European PURE-MIND biomarker participants were used for observational association analyses.

We sought replication of our observational findings in GS ^[34, 35]^, which was recruited through re-contact of the Generation Scotland: Scottish Family Health Study (GS:SFHS) ^[36, 37]^. GS:SFHS is a population- and family-based cohort of >24,000 individuals from Scotland. GS:SFHS participants were recruited between 2006 and 2011. Upon recruitment, participants attended a clinic where detailed health, cognitive, and lifestyle information, and biological samples were collected. Between 2015 and 2018, a subset of the GS:SFHS participants completed additional health and cognitive assessments, brain MRI, and provided blood samples for proteomic analysis. Up to 1053 GS participants were available for replication analyses.

### Ethical approval

All centres contributing to PURE were required to obtain approval from their respective ethics committees (Institutional Review Boards). Participant data is confidential and only authorized individuals can access study-related documents. The participants’ identities are protected in documents transmitted to the Coordinating Office, as well as biomarker and genetic data. Participants provided informed consent to obtain baseline information, and to collect and store genetic and other biological specimens.

The GS:SFHS obtained ethical approval from the NHS Tayside Committee on Medical Research Ethics, on behalf of the National Health Service (reference: 05/S1401/89). All participants provided broad and enduring written informed consent for biomedical research. GS:SFHS has Research Tissue Bank Status (reference: 15/ES/0040), providing generic ethical approval for a wide range of uses within medical research. The imaging subsample of GS:SFHS (referred to as “GS” herein) received ethical approval from the NHS Tayside committee on research ethics (reference 14/SS/0039). All experimental methods were in accordance with the Helsinki declaration.

### Assessment of between-cohort differences

Between cohort differences in quantitative variables (age, DSST score, BMI, blood pressure, and brain MRI volumes) were assessed using two-sample t-tests. Categorical variables (sex, education type, and disease status) where the smallest count in any cell of the contingency table was greater than five were assessed using a chi-squared test; otherwise, a Fisher’s Exact Test was employed. Statistical significance was defined as *p* < 0.05.

### Assessment of cognitive function

General cognitive ability was measured in PURE-MIND and GS by trained assessors using the DSST (Wechsler Adult Intelligence Scale, 3rd Edition) ^[38]^. The DSST is a pencil and paper test in which participants must match symbols to numbers according to a key. Participants were scored according to the number of correct matches made within two minutes (maximum score: 133). The DSST measures several cognitive functions, including associative learning and executive function ^[39]^, and DSST performance is highly correlated with the general intelligence factor, *g*. PURE-MIND participants completed the Montreal Cognitive Assessment (MoCA) ^[11]^, a questionnaire-based test with scores 0 to 30. A score of 26 or higher is considered normal ^[11]^.

### Measurement of plasma protein expression

In the PURE biomarker cohort, 1196 plasma protein levels were measured by proximity extension assay using the Olink Proseek Target 96 reagent kit (Olink, Uppsala, Sweden) in 12066 participants (including 3735 European, 4695 Latin, and 1436 Persian). Following pre-processing and quality control steps (**Supplementary Information**), measurements were available for 1160 biomarkers in 8369-9154 European, Latin, or Persian participants (depending on biomarker-specific missingness).

In GS, plasma protein levels were measured with the SOMAscan assay platform (SomaLogic Inc.), as described previously ^[40]^. Following initial data processing and quality control steps, measures of 4058 proteins were available in 1095 participants. Prior to analysis, protein abundance measurements were log-transformed and rank-based inverse normalised.

### Brain imaging

PURE-MIND participants enrolled in the PURE biomarker cohort were scanned at four sites in Canada (three at 1.5T (two on General Electric (GE) scanners, one on a Phillips scanner), one at 3T (GE)). The brain imaging phenotypes assessed in this study were total brain volume (excluding ventricles), total white matter volume, hippocampal volume, average cortical thickness, a multi-region composite thickness measure designed to differentiate Alzheimer’s disease patients from clinically normal participants ^[41]^, silent brain infarcts (SBI), cerebral microbleeds (CMB), and WMH volumes. These will henceforth be referred to as the “structural brain phenotypes”. Further information about the derivation of the structural brain phenotypes is available in the **Supplementary Information**.

### Genotyping and imputation of PURE-MIND

PURE participant genotypes (Thermofisher Axiom Precision Medicine Research Array r.3) were called using Axiom Power Tools and in-house scripts. Quality control steps are described in the **Supplementary Information**.

Imputation was performed on the 749,783 genotyped variants following the TOPMed Imputation server pipeline (https://imputation.biodatacatalyst.nhlbi.nih.gov/). Further details are in the **Supplementary Information**.

### Assessment of the association between protein biomarkers and cognitive function and structural imaging phenotypes

We assessed the association between standardised protein levels and cognitive and structural brain phenotypes using two-tailed linear (DSST, MoCA, total brain volume, white matter volume, hippocampal volume, WMH volume, cortical thickness) or logistic (CMB, SBI) regression. The cognitive or structural brain phenotype-of-interest was the dependent variable with the standardised protein expression level, age, age^2^, sex, and the first ten genetic principal components as independent variables. A sensitivity analysis was performed for DSST-associated proteins in which we further adjusted for education (a categorical variable with levels: (i) no education; (ii) high school or less; (iii) trade school; and (iv) college or university). We calculated Pearson’s correlation coefficient to assess the pairwise correlations between DSST-associated proteins. Within each analysis, we applied a Bonferroni correction to determine statistical significance, yielding the following significance thresholds: *p* < 4.31 x 10^−5^ when assessing associations with 1160 proteins; *p* < 2.5 x 10^−3^ when assessing associations with the five DSST-associated proteins across four DSST-associated structural brain phenotypes; and *p* < 5 x 10^−3^ when assessing 10 pairwise correlations between proteins.

We performed replication analyses in GS for the significant proteins identified in PURE-MIND. Two-tailed mixed effects models were fitted using the lmekin function from the R package coxme v.2.2.17^[42]^ to assess the association of the outcome variable (DSST performance, total brain volume (excluding ventricles), cerebral white matter volume, hippocampal volume, and WMH volume) with standardised protein expression, covarying for age, age^2^, sex, study site (Dundee or Aberdeen), the delay between blood sampling and protein extraction, depression (a binary variable representing lifetime depression status), and a kinship matrix. When a brain volume phenotype was the outcome variable, additional covariates were included to account for ICV, the interaction between ICV and study site (to account for a site-associated batch effect on ICV measurement), and whether there was manual intervention using tools within Freesurfer during the quality control process. Replication was defined as a concordant direction of effect, meeting a Bonferroni-corrected threshold of *p* < 1.67 x 10^−2^ (accounting for the assessment of three DSST-associated proteins) or *p* < 7.14 x 10^−3^ (accounting for the assessment of seven structural brain phenotype-protein combinations).

### Assessment of the association of DSST performance with MRI-derived structural brain phenotypes

To identify mediators of the association between protein expression and DSST performance, we first established the structural brain phenotypes that satisfied the requirements of potential mediators (i.e. associated with both DSST performance and at least one DSST-associated protein), and then formally tested the meditation relationship by bootstrap mediation analyses.

We estimated the association between DSST performance and structural brain phenotypes in PURE-MIND using linear models. All brain volume measurements were normalised to ICV and the models included covariates for age, age^2^, sex, and the first ten genetic principal components. We defined statistical significance as *p* < 0.00625 (Bonferroni correction for eight phenotypes; two-tailed) and sought replication of significant associations (N = 5) in GS. In GS, brain volumes were residualised for ICV, scanner location, the interaction between ICV and scanner location, and whether there was manual intervention during the quality control process. The resultant residuals were included as the dependent variable in a mixed effects model with DSST score, age, age^2^, sex, depression, and a kinship matrix as independent variables. Statistical significance was defined as *p* < 0.0125 (two-tailed).

The DSST performance-associated brain MRI phenotypes (N = 4) were assessed as potential mediators of the protein level-DSST associations (N = 3, yielding a total of N = 9 mediations to assess) using bootstrap mediation analysis in PURE-MIND. Analyses were performed using the R package “mediation” ^[43]^ with 1000 bootstraps. We corrected for the nine potential mediation relationships assessed using a Bonferroni-corrected threshold of *p* < 5.56 x 10^−3^.

### Functional and tissue-specific expression enrichment analyses

Proteins associated with DSST performance at *p* < 0.05 in PURE-MIND were included in functional and tissue-specific expression analyses in three groups: (i) all proteins; (ii) positively associated proteins; and (iii) negatively associated proteins. Enrichment was assessed relative to all proteins in our dataset that passed quality control (N =1160). Functional enrichment analyses were performed using WebGestalt (http://www.webgestalt.org/) ^[44]^ using default parameter settings for the over-representation analysis method to assess enrichment for: (i) gene ontology categories (biological processes, molecular functions, and cellular compartments); (ii) Reactome pathways; and (iii) disease-associated genes (Disgenet). Tissue-specific enrichment analyses were performed using the “GTEx v8: 54 tissue types” and “GTEx v8: 30 general tissue types” gene expression datasets in FUnctional Mapping and Annotation (FUMA) ^[45]^. For both the functional enrichment and tissue expression analyses, enrichment was assessed using a hypergeometric test and significant enrichment was defined as a Benjamini-Hochberg-adjusted *p* < 0.05, correcting for the number of tests performed within each analysis platform. Analyses were performed using web interfaces accessed on 18/04/2022 (WebGestalt and FUMA) and 14/01/2023 (FUMA).

### Two-sample forward MR analyses

We performed two-sample forward MR analyses to identify potentially causal associations between genetically predicted plasma protein levels and: (i) cognitive function; (ii) structural brain phenotypes (total brain volume, cerebral white matter volume, hippocampal volume, WMH volume, and CMB); and (iii) disease outcomes (Alzheimer’s disease, all stroke, stroke subtypes (ischaemic, cardioembolic, large artery, and small vessel), and intracranial aneurysm).

Associations between single nucleotide polymorphisms (SNPs) and plasma protein expression levels were calculated in PURE (**Supplemental Information**). Following quality control, a set of pQTLs that was independent (r^2^ < 0.1) in all three populations was retained. Sensitivity analyses were performed in which the pruning threshold was adjusted to r^2^ < 0.01.

The independent set of pQTLs were assessed for their associations with cognitive function, structural brain phenotypes, and disease outcomes using summary statistics from published studies ^[46–53]^.

MR analyses were performed using the R packages MRBase for TwoSample MR v.0.5.6 ^[54]^, mr.raps v.0.4.1 ^[55]^, and MRPRESSO v.1.0 ^[56]^. We employed several complementary MR approaches: IVW ^[57]^, weighted median ^[58]^, robust adjusted profile scores (RAPS) ^[55]^, MR-Egger ^[59]^, and MR-PRESSO ^[56]^. We adopted the IVW approach as our primary methodology and defined statistical significance using a liberal within-outcome variable Bonferroni correction for the proteins (CA14, CDCP1, and MOG) that could be assessed, yielding a significance threshold of *p* < 0.0167 (or *p* < 0.025 or 0.05 when an outcome could only be assessed for two or one protein(s)). Further details of the MR analyses are included in the **Supplementary Information**.

### Pairwise Conditional Analysis and Co-localisation Analysis (PWCoCo)

PWCoCo ^[31, 60]^ was performed to assess the existence of a shared causal variant between (i) pQTLs for each of the five proteins-of-interest and (ii) variants associated with the outcomes assessed in the two-sample MR analyses (**Supplementary Information**).

### Software

Statistical analyses and plot generation were performed in R (versions 3.6.0, 4.1.1, 4.1.2, 4.2.0, 4.2.1).

## Data availability

The terms of consent for PURE participants preclude the sharing of individual-level data. Individual level data is available through collaboration with PURE researchers (https://www.phri.ca/research/pure/). Summary-statistics for the analyses presented here are available in the supplementary materials.

According to the terms of consent for GS participants, applications for individual-level data must be reviewed by the GS Access Committee. Complete summary statistics are available in the supplementary materials for the protein-DSST score associations assessed in this study.

## Code availability

The code used to generate the results in this study is available on reasonable request from the corresponding author.

## Funding and acknowledgements

### PURE

The PURE study is an investigator-initiated study that is funded by the Population Health Research Institute, the Canadian Institutes of Health Research (CIHR), Heart and Stroke Foundation of Ontario, support from CIHR’s Strategy for Patient Oriented Research, through the Ontario SPOR Support Unit, as well as the Ontario Ministry of Health and Long-Term Care and through unrestricted grants from several pharmaceutical companies (with major contributions from AstraZeneca [Canada], Sanofi-Aventis [France and Canada], Boehringer Ingelheim [Germany and Canada], Servier, and GlaxoSmithKline), and additional contributions from Novartis and King Pharma and from various national or local organisations in participating countries as follows: Argentina—Fundacion ECLA; Bangladesh—Independent University, Bangladesh and Mitra and Associates; Brazil—Unilever Health Institute, Brazil; Canada—Public Health Agency of Canada and Champlain Cardiovascular Disease Prevention Network; Chile—Universidad de la Frontera; Colombia—Colciencias (grant number 6566– 04–18062); South Africa—The North-West University, SANPAD (SA and Netherlands Programme for Alternative Development), National Research Foundation, Medical Research Council of South Africa, The South Africa Sugar Association, Faculty of Community and Health Sciences; Sweden—grants from the Swedish State under the Agreement concerning research and education of doctors, the Swedish Heart and Lung Foundation, the Swedish Research Council, the Swedish Council for Health, Working Life and Welfare, King Gustaf V’s and Queen Victoria Freemasons Foundation, AFA Insurance, Swedish Council for Working Life and Social Research, Swedish Research Council for Environment, Agricultural Sciences and Spatial Planning, grant from the Swedish State under (LäkarUtbildningsAvtalet) Agreement, and grant from the Västra Götaland Region; and United Arab Emirates—Sheikh Hamdan Bin Rashid Al Maktoum Award for Medical Sciences, Dubai Health Authority, Dubai. The PURE biomarker project was supported by Bayer and the CIHR. The biomarker project was led by PURE investigators at the Population Health Research Institute (Hamilton, Canada) in collaboration with Bayer scientists. Bayer directly compensated the Population Health Research Institute for measurement of the biomarker panels, scientific, methodological, and statistical work. Genetic analyses were supported by CIHR (G-18–0022359) and Heart and Stroke Foundation of Canada (application number 399497) in the form of funding to GP.

### GS

This work was supported by the Wellcome Trust [104036/Z/14/Z, 220857/Z/20/Z, and 216767/Z/19/Z] and an MRC Mental Health Data Pathfinder Grant [MC_PC_17209] to AMM. DAG is funded by the Wellcome Trust Translational Neuroscience PhD Programme at the University of Edinburgh [108890/Z/15/Z]. LS and ANH are supported by Medical Research Council [MR/L023784/2]: Dementias Platform UK. LS is also supported by a Medical Research Council Award to the University of Oxford [MC_PC_17215]. AS is supported through the Wellcome-University of Edinburgh Institutional Strategic Support Fund (Reference 204804/Z/16/Z), and indirectly through the Lister Institute of Preventive Medicine award with reference 173096. Generation Scotland received core support from the Chief Scientist Office of the Scottish Government Health Directorates [CZD/16/6] and the Scottish Funding Council [HR03006]. Genotyping of the GS:SFHS samples was carried out by the Genetics Core Laboratory at the Clinical Research Facility, Edinburgh, Scotland, and was funded by the UK’s Medical Research Council and the Wellcome Trust [104036/Z/14/Z].

We are grateful to all the families who took part in Generation Scotland, the general practitioners and the Scottish School of Primary Care for their help in recruiting them, and the whole Generation Scotland team, which includes interviewers, computer and laboratory technicians, clerical workers, research scientists, volunteers, managers, receptionists, healthcare assistants and nurses.

We thank Dr Alison Offer for assistance in producing the forest plots.

