## Supplementary table legends for "The circulating proteome and brain health: Mendelian randomisation and cross-sectional analyses"

**Supplementary table 1.** Observational analyses of the associations between the levels of 1160 plasma proteins and digit symbol substitution test (DSST) performance in PURE-MIND. Results are shown for the primary analytic approach and a sensitivity analysis in which education was adjusted for. Protein levels were deemed to be associated with DSST performance when *p* < 4.31 x 10^-5^.

**Supplementary table 2.** Assessment of the correlation between proteins whose expression is significantly correlated with DSST performance in PURE-MIND. Pearson's correlation coefficients together with 95% confidence intervals are shown in the upper triangle, whilst *p*-values are shown in the lower triangle. Significant correlations (*p* < 0.005) are indicated by bold text.

**Supplementary table 3.** Observational analyses of the associations between the levels of 1160 plasma proteins and Montreal Cognitive Assessment (MoCA) performance in PURE-MIND. Protein levels were deemed to be associated with MoCA performance when *p* < 4.31 x 10^-5^.

**Supplementary table 4.** Summary statistics from PURE-MIND and GS for proteins whose levels were nominally significantly (*p* < 0.05) associated with DSST in PURE-MIND.

**Supplementary table 5.** Enrichment analyses for gene sets showing tissue-specific mRNA expression patterns. Hypergeometric tests were performed to assess the enrichment or depletion of sets of genes expressed in a tissue-specific manner amongst those proteins showing nominally significant (*p* < 0.05) association with DSST performance in PURE-MIND. Separate analyses were performed for all nominally significant proteins (“All”), and nominally significant proteins that showed positive (“Up”) and negative (“Down”) associations with DSST performance. For each gene set, the number of genes in the set (“N genes”), the number of set members in the target list (“N overlap”), *p*-value and false discovery rate (FDR)-corrected *p*-value are shown. Significant enrichment was defined as FDR-corrected *p* < 0.05.

**Supplementary table 6.** Enrichment analyses for gene ontology categories (biological processes, molecular functions, and cellular compartments), Reactome pathways, and disease associations amongst proteins showing nominally significant (*p* < 0.05) association with DSST performance in PURE-MIND. Separate analyses were performed for all nominally significant proteins (“All”), and nominally significant proteins that showed positive (“Up”) and negative (“Down”) associations with DSST performance. For each set of proteins (all, down up), the ten most significantly enriched gene sets are shown. For each gene set, the total number of genes in that set (“Size”), the expected number of set members in the target list (“Expected”), the ratio of the observed to the expected number of set members in the target list (“Ratio”), and the hypergeometric test *p*-value and false discovery rate (FDR)-corrected *p*-value are shown. Significant enrichment was defined as FDR-corrected *p* < 0.05.

**Supplementary table 7.** Observational analyses of the associations between MRI-derived structural brain phenotypes and performance on the DSST in PURE-MIND and GS. Volumetric phenotypes were corrected for intracranial volume. Phenotypes identified as being significantly associated in PURE-MIND (*p* < 0.00625) were assessed for replication in GS, in which replication was defined as *p* < 0.0125.

*composite measure comprising regions selected for discrimination of Alzheimer’s disease cases from controls (Schwarz et al., 2016)

**Supplementary table 8.** Assessment of MRI-derived structural brain phenotypes as mediators of the association between plasma protein levels and DSST performance. Mediation was assessed by bootstrap mediation analysis (1000 resamples) and the average causal mediation effect (ACME) and proportion of the total effect attributable to the mediator calculated. Significant mediation was defined as an ACME *p* < 0.05/number proteins assessed for a given brain phenotype.

**Supplementary table 9.** Significant *cis*-pQTLs for CA14, CDCP1, and MOG through a fixed effects meta-analysis of the European, Latin, and Persian populations (*p* < 5 x 10^-^6). Summary statistics from the meta-analysis are shown, together with summary statistics from the individual population association analyses. For each SNP, its chromosomal location, and reference and alternative alleles are shown.

**Supplementary table 10.** Two-sample forward MR analyses to detect potentially causal effects of variation in plasma levels of five DSST performance-associated proteins on cognitive function, structural brain phenotypes, and stroke, intracranial aneurysm, and Alzheimer’s disease. Summary statistics are shown for each of four MR methodologies (inverse variance weighted (IVW), weighted median, robust adjusted profile score (RAPS), and MR-Egger), together with the number of SNPs included as IVs for protein level, the proportion of variance in protein level explained by these SNPs, and the F-statistic for the association between the IVs and protein levels. The Egger intercept test *p*-value and Cochran’s Q-test *p*-value are shown to allow the assessment of directional and horizontal pleiotropy, respectively. The correct causal direction was defined as the IVs explaining a larger proportion of the variance in the exposure than the outcome, and Steiger’s test was performed to statistically assess whether the correct causal direction had been tested. NB. A lack of IVs precluded two-sample MR analyses for small vessel stroke and white matter hyperintensity volume. Sensitivity analyses were performed in which the linkage disequilibrium threshold used to prune SNPs was adjusted from r^2^ < 0.1 to r^2^ < 0.01. The results of the sensitivity analyses are presented alongside the results of the primary analyses. Random effects meta-analyses were also performed for those associations attaining statistical significance in our primary analyses. Where the results from the primary analysis met our criteria for significance (IVW p-value < 0.05/number proteins for which the outcome was assessed, no evidence of pleiotropy, and corroboration of the direction of effect from at least two other MR approaches), yellow highlighting indicates the relevant rows.
