## Supplementary Information for "The circulating proteome and brain health: Mendelian randomisation and cross-sectional analyses"

1. **Supplementary figures**

**A**

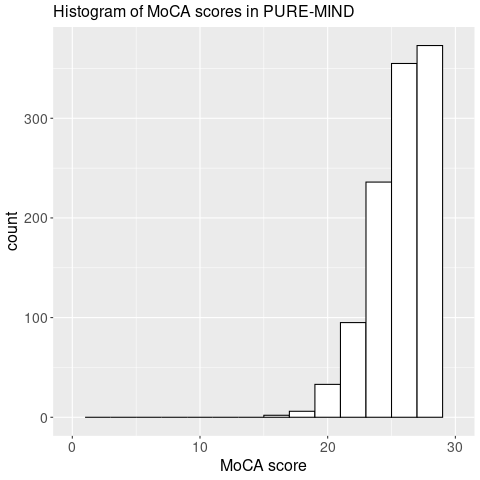

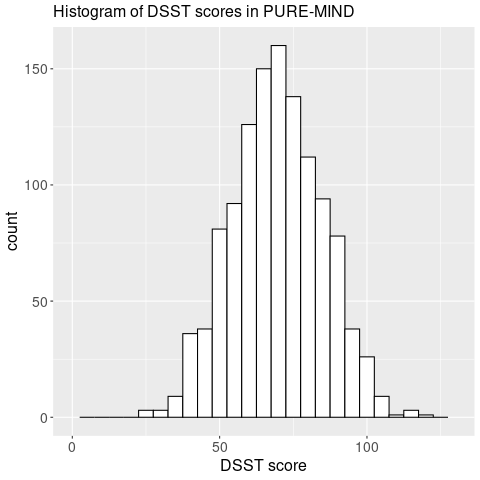

**B**

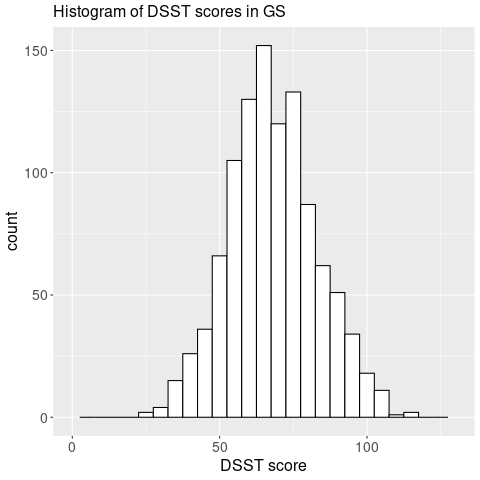

**C**

**Supplementary figure 1.** Histograms to show the distribution of DSST scores in PURE-MIND (A), MoCA scores in PURE-MIND (B), and DSST scores in GS (C). The x-axes indicate cognitive test score, and the y-axes indicate frequency.

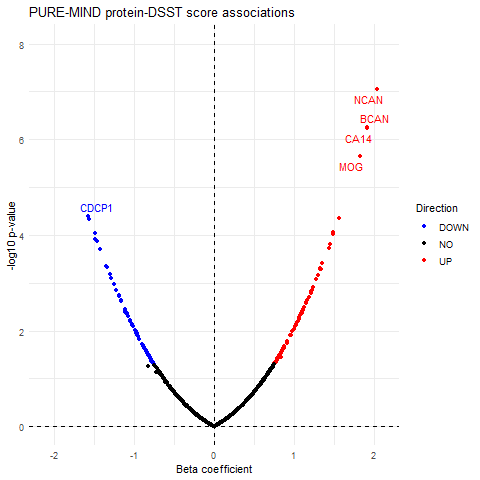

**Supplementary figure 2.** Volcano plot showing the beta-coefficient (x-axis) and -log_10_ *p*-value (y-axis) of the associations between 1160 plasma proteins and DSST score in PURE-MIND. Each point represents a single protein, and points are colour-coded to indicate whether they showed no association (black), or a nominally significant (*p* < 0.05) negative (blue) or positive (red) association with DSST score. Proteins that attained a Bonferroni-corrected threshold for statistical significance (*p* < 4.31 x 10^-5^) are labelled with their protein symbol.

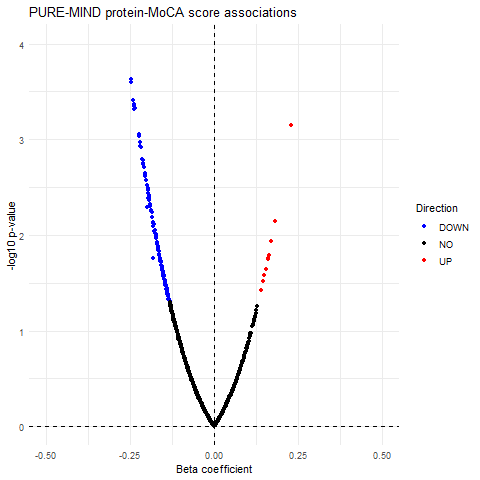

**Supplementary figure 3.** Volcano plot showing the beta-coefficient (x-axis) and -log_10_ *p*-value (y-axis) of the associations between 1160 plasma proteins and MoCA score in PURE-MIND. Each point represents a single protein, and points are colour-coded to indicate whether they showed no association (black), or a nominally significant (*p* < 0.05) negative (blue) or positive (red) association with DSST score.

2. **Supplementary methods**

**The PURE biomarker study**

The PURE biomarker study is a nested case-cohort study of the original PURE study ^[1]^ with protein biomarkers and genotyping data ^[2]^. Cases were selected if they had experienced at least one of the following adverse health events: myocardial infarction, stroke, heart failure, type II diabetes, or death from any cause. Cohort members were selected by random sampling to obtain a group of participants who were frequency-matched by major country-specific ethnicity to the cases.

**PURE-MIND**

Participants from selected countries in PURE ^[1]^ were invited to participate in PURE-MIND if they were aged ≥ 39 years, had no history of stroke, dementia, or other neurological disease; had no contraindication to MRI; and could complete cognitive assessments ^[3]^.

**Plasma protein measurement in the PURE biomarker study**

Proteomic and genetic analyses were conducted in the Clinical Research Laboratory & Biobank – Genetic & Molecular Epidemiology Laboratory (CRLB-GMEL), Hamilton, Canada. In the PURE biomarker cohort, plasma protein levels were measured by proximity extension assay using the Olink Proseek Target 96 reagent kit (Olink, Uppsala, Sweden). Thirteen panels (Cardiometabolic; Cardiovascular Disease II and III; Cell Regulation; Development; Immune Response; Inflammation, Metabolism; Neuro Exploratory; Neurology; Oncology I and III; and Organ Damage) were used to measure a total of 1196 biomarkers in 12066 participants, of which (3735 European, 4695 were Latin, and 1436 were Persian). The analytical performance of these panels has been validated previously and further information can be found elsewhere (<https://www.olink.com/products-services/target/>).

Quality control and pre-processing of the PURE biomarker study protein measurements were performed as described previously for the Cardiovascular Disease II panel ^[2]^, with the exception that the data were quantile normalised within three, rather than two, reagent lots. Missing biomarker values were imputed by the mean, separately for each reagent lot, and all values were rank-based inverse normalised by reagent lot, sex and ethnic group. Where multiple biomarker measurements from different proximity extension assays were available for a single protein, the mean value was taken.

**Genotyping and imputation of PURE-MIND**

Samples were removed if: they had a low signal-to-noise contrast (Dish Quality Control < 0.82); low quality control rate (QCCR < 0.97); <95% call rate; disagreement between self-reported sex and/or ethnicity and genetically determined sex and/or ethnicity; were duplicated; or had excess heterozygosity. We removed variants with: a call rate <98.5%; Hardy-Weinberg equilibrium *p* < 1 x 10^-5^; plate or batch effects; non-Mendelian segregation within families; and/or a minor allele frequency <0.005%. Following quality control, 749,783 variants remained ^[2]^.

Imputation was performed on the 749,783 genotyped variants following the TOPMed Imputation server pipeline (<https://imputation.biodatacatalyst.nhlbi.nih.gov/>), using the TOPMed release 2 reference panel ^[4]^. EAGLE v2.4 ^[5]^ and Minimac4 programs were applied for phasing and imputation, respectively. Imputed variants with an info score ≥ 0.3 and MAF ≥ 0.01, which did not deviate from Hardy-Weinberg equilibrium (*p* ≥ 1 x 10^-5^) were retained.

**Identification of pQTLs in PURE**

Associations between single nucleotide polymorphisms (SNPs) and plasma protein expression levels were calculated in PURE. SNPs located within 200 kilobases up- or downstream of the RefSeq transcript corresponding to a protein-of-interest were assessed as potential pQTLs (*p* < 5 x 10^-6^) through inverse variance-weighted fixed effects meta-analysis using METAL ^[6]^ of the European (N = 3514), Latin (N = 4309), and Persian (N = 1332) populations. Missense variants and SNPs affecting splice sites, together with variants in LD (r^2^ ≥ 0.9) with them, were excluded in order to avoid effects on antibody binding. The pQTL model has been described previously ^[2]^. An independent set of pQTLs obtained by pruning (r^2^ < 0.1) within each of the European, Latin, and Persian subgroups separately, using LD information derived from each separate population subgroup genotype data within PURE using Plink v1.90b3o. The retained overlapping pQTLs were independent in all three populations. Sensitivity analyses were performed in which the pruning threshold was adjusted to r^2^ < 0.01.

**Two-sample MR**

The IVW approach as our primary MR methodology, as it has the greatest statistical power; however, it also makes the most assumptions. Hence, we reported IVW findings only where there was: (i) no evidence of pleiotropy; and (ii) corroboration of the direction of effect from at least two other MR approaches. An MR-Egger intercept *p* < 0.05 was deemed to indicate directional pleiotropy. Heterogeneity amongst instrumental variables, suggestive of horizontal pleiotropy, was indicated by a significant Cochran’s Q (*p* < 0.05). If Cochran’s Q was significant, MR-PRESSO ^[7]^ was performed, and, if the MR-PRESSO global test was significant (*p* < 0.05), MR-PRESSO with outlier removal was performed. In addition to the above conditions, we only reported results where there were at least three IVs, there was no evidence of weak instrument bias (*F*-statistic > 10) ^[8]^, and when the correction causal direction had been assessed (indicated by the instrumental variables explaining a greater proportion of the variance in the exposure than in the outcome, and a Steiger test *p* < 0.05). For the sensitivity analyses, in which a more stringent r^2^ threshold was used to select independent pQTLs, only one or two IVs were available for each analysis. When two IVs were available, results from the IVW approach are reported, and when one IV was available, results from the Wald ratio test were reported. Statistical significance was defined as *p* < 0.05.

To assess whether our MR results had been affected by between-population heterogeneity in pQTL effect estimates, we carried out MR analyses in each of the three populations separately (using population-specific pQTL effect estimates), and combined the MR effect estimates by random effects meta-analysis using metafor (version 3.0-2). Where more than one IV was available in all three populations, the meta-analysed IVW results are presented as the primary results, and the meta-analysed weighted median and MR-Egger results are presented as secondary approaches. Where only one IV was available in at least one of the populations, the meta-analysed Wald ratio results are presented. Statistical significance was defined as *p* < 0.05.
